# Detection of hypertrophic cardiomyopathy on electrocardiogram using artificial intelligence

**DOI:** 10.1101/2024.11.19.24317545

**Authors:** James M Hillis, Bernardo C Bizzo, Sarah F Mercaldo, Ankita Ghatak, Ashley L MacDonald, Madeleine A Halle, Alexander S Schultz, Eric L’Italien, Victor Tam, Nicole K Bart, Filipe A Moura, Amine M Awad, David Bargiela, Sarajune Dagen, Danielle Toland, Alexander J Blood, David A Gross, Karola S Jering, Mathew S Lopes, Nicholas A Marston, Victor D Nauffal, Keith J Dreyer, Benjamin M Scirica, Carolyn Y Ho

## Abstract

**Background:** Hypertrophic cardiomyopathy (HCM) is associated with significant morbidity and mortality including sudden cardiac death in the young. Its prevalence is estimated to be 1 in 500, although many people are undiagnosed. The ability to screen electrocardiograms (ECGs) for its presence could improve detection and enable earlier diagnosis.

**Objectives:** This study evaluated the accuracy of an artificial intelligence device (Viz HCM) in detecting HCM based on 12-lead ECG.

**Methods:** The device was previously trained using deep learning and provides a binary outcome (HCM suspected or not suspected). This study included 293 HCM-Positive and 2912 HCM-Negative cases, which were selected from three hospitals based on chart review incorporating billing diagnostic codes, cardiac imaging, and ECG features. The device produced an output for 291 (99.3%) HCM-Positive and 2905 (99.8%) HCM-Negative cases.

**Results:** The device identified HCM with sensitivity 68.4% (95% CI: 62.8-73.5%), specificity 99.1% (95% CI: 98.7-99.4%) and area under the curve 0.975 (95% CI: 0.965-0.982). With assumed population prevalence of 0.002 (1 in 500), the positive predictive value was 13.7% (95% CI: 10.1-19.9%) and the negative predictive value was 99.9% (95% CI: 99.9-99.9%). The device demonstrated consistent performance across demographic and technical subgroups.

**Conclusions:** The device identified HCM based on 12-lead ECG with good performance. Coupled with clinical expertise, it has the potential to augment HCM detection and diagnosis.

## Introduction

Hypertrophic cardiomyopathy (HCM) is a complex myocardial disorder that may result in substantial morbidity and mortality, including advanced heart failure and sudden cardiac death (1). Its prevalence is estimated to be 1 in 500, although many people are undiagnosed (2–6). The ability to screen electrocardiograms (ECGs) for its presence could improve detection and enable earlier diagnosis. Given HCM is often genetic, this approach has the potential to enhance the care of both individual patients and families.

A key challenge with diagnosing HCM is that it relies on a dichotomous measure of left ventricular wall thickness (threshold slightly modified based on family history) after excluding other potential factors that may increase wall thickness, particularly hypertension and storage or infiltrative disorders (7). ECGs may show findings consistent with HCM, such as left ventricular hypertrophy, although such findings are not specific to HCM (8). It has been proposed that an ECG artificial intelligence (AI) algorithm may be able to detect features that are more specific for HCM (9). Such an algorithm could facilitate screening patients for HCM, triggering further clinical evaluation when appropriate, to confirm the diagnosis.

This study investigated the Viz HCM software-as-a-medical-device, which utilizes ECG as its input and has subsequently been granted de novo approval through the United States Food and Drug Administration (FDA) (10). It aimed to evaluate the performance of the device by comparing its outputs to ground truth interpretations as determined by expert clinical review. To assess robustness and generalizability, it also explored the performance in subgroups based on patient demographics, technical parameters, potentially confounding ECG findings and co-existing diagnoses (including mimics of HCM).

## Methods

### Study design

This retrospective standalone model performance study was conducted using ECG cases from three hospitals within the Mass General Brigham (MGB) network between July 1, 2017, and June 30, 2022. These three hospitals had not provided any training data for the model. The study was approved by the MGB Institutional Review Board with waiver of informed consent. It was conducted in accordance with relevant guidelines and regulations including the Health Insurance Portability and Accountability Act (HIPAA). This report followed the Standards for Reporting Diagnostic Accuracy (STARD 2015) reporting guideline.

### Cohort selection process

This study included 300 HCM-Positive and 3000 HCM-Negative ECG cases from distinct patients. These cases were identified from a list of candidate cases in a consecutive manner until the required cohort size was achieved. The cohort selection occurred through a clinical record search based on ICD-9 and/or ICD-10 codes that was confirmed through a manual chart review. The cohort was essentially finalized after these processes with a small number of cases being excluded during the subsequent ECG annotations and model inference (Figure 1).

**Figure 1.**
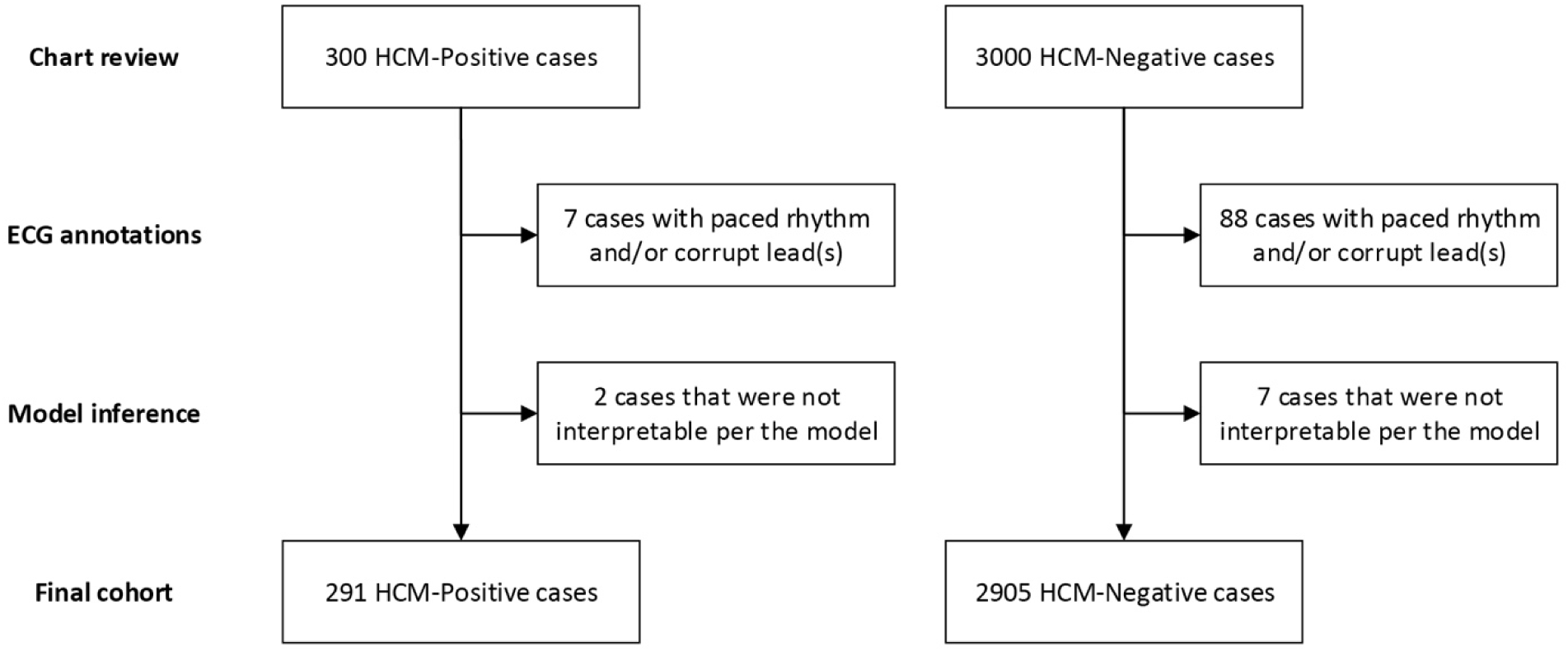
Cohort selection flowchart.

The clinical record search for HCM-Positive cases identified patients with the ICD-9 and/or ICD-10 codes for HCM (ICD-9: 425.1 Hypertrophic cardiomyopathy; ICD-10: I42.1 Obstructive hypertrophic cardiomyopathy or I42.2 Other hypertrophic cardiomyopathy) through the Research Patient Data Registry (11) at MGB. The clinical record search for HCM-Negative cases identified patients without these ICD-9 and/or ICD-10 codes; given the large number of available ECG cases to consider for the HCM-Negative cases, ECGs were considered from five random months for each hospital (this strategy amounted to 200 cases for each hospital from each month). Both searches also excluded patients if they had an ICD-9 and/or ICD-10 code for a pacemaker (ICD-9: V45.01; ICD-10: Z95.0), which was an exclusion criterion given the alteration of ECGs in the presence of paced rhythms. There was no restriction on patient setting within the hospital (i.e., ECGs were considered from emergency patients, inpatients and outpatients).

The identified cases were then placed in a randomized order such that confirmatory chart reviews could follow the principles of consecutive cohort selection based on this order until sufficient cases were selected. These chart reviews were performed by board-certified cardiologists who were assisted by resident physicians and critical care nurses for components not requiring cardiology knowledge (e.g., a patient not having had an imaging study); all HCM-Positive cases and ∼55% of HCM-Negative cases included in the final cohort were reviewed by a board-certified cardiologist. This information was entered directly into the REDCap electronic data capture tool hosted at MGB (12,13). Once the HCM-Positive and HCM-Negative cohorts were finalized, the ECG cases underwent deidentification for use in subsequent steps.

### HCM-Positive case chart review

A manual chart review confirmed the patient’s diagnosis of HCM including whether it was obstructive or non-obstructive HCM. The ECG used for analysis was selected by taking the first ECG of the day on the date closest to the first echocardiography and/or cardiac magnetic resonance imaging (CMR) study that established the diagnosis of HCM. Cases were excluded if the patient was <18 years of age, had no confirmatory echocardiography and/or CMR, had an alternate diagnosis to HCM, had received septal myectomy or alcohol ablation prior to the ECG, had no evidence of left ventricular hypertrophy by both the Cornell and Sokolow-Lyon voltage criteria (i.e., cases were included if either criterion was met), or had pacemaker presence and/or corrupt lead(s). The results of genetic testing, left ventricular wall thickness and left ventricular ejection fraction were also obtained from each patient’s chart when available; the left ventricular wall thickness was based on the reported intraventricular septum thickness on echocardiography or the maximal wall thickness when an apical variant; the left ventricular wall thickness was based on the reported maximal wall thickness on CMR.

### HCM-Negative case chart review

A manual chart review confirmed the patient’s absence of a diagnosis of HCM and identified the ECG used for analysis, which was the first one within the selected month at that hospital. The initial chart review involved searching for the terms “HCM” and “hypertrophic cardiomyopathy”, determining if the patient had had an echocardiography and/or CMR study within the study date range, and ensuring the selected ECG did not have pacemaker presence and/or corrupt lead(s). If any of these criteria were met then a cardiologist reviewed the case to ensure the patient did not have a diagnosis of HCM, the ECG did not have pacemaker presence and the ECG did not have corrupt lead(s) before it was included in the final cohort. If none of the exclusion criteria were met, the case was included in the final cohort.

### ECG annotations

The ECG cases were all then reviewed by a cardiologist for the presence of 16 different findings or abnormalities: pacemaker presence, atrial fibrillation, premature atrial contraction(s) / premature ventricular contraction(s), left axis deviation, atrial enlargement, left ventricular hypertrophy, right ventricular hypertrophy, increased precordial voltages, acute ischemic change(s), myocardial infarct of indeterminate age, non-specific ST segment change(s), non-specific T wave abnormality, ECG contains a corrupt lead(s), other ECG artifact or noise, other pathologic finding (with free text), and no pathologic finding. This step incorporated mixed HCM-Positive and HCM-Negative cases such that the ECG annotations would not be influenced by the knowledge of a case being positive or negative. If an ECG was annotated as having pacemaker presence or containing a corrupt lead(s) then it was excluded. These annotations were performed using the FDA-cleared eUnity image visualization software (Version 6 or higher) and a previously described internal web-based annotation system that utilized REDCap (12–14).

### Comorbid conditions

The clinical record search also returned information about the presence of comorbid conditions based on ICD-9 and ICD-10 codes. The presence or absence of the following cardiopulmonary conditions was recorded: ischemic heart disease (ICD-9: 410-414; ICD-10: I20-I25), pulmonary hypertension (ICD-9: 416.0; ICD-10: I27.0, I27.2), pulmonary heart disease (ICD-9: 416.8, 416.9; ICD-10: I27.8, I27.9), nonrheumatic aortic stenosis (ICD-9: 424.1; ICD-10: I35.0, I35.2), left bundle branch block, unspecified (ICD-9: 426.2, 426.3; ICD-10: I44.7), right bundle branch block (ICD-9: 426.4, 426.5; ICD-10: I45.0, I45.1, I45.2, I45.3), pre-excitation syndrome (which incorporates Wolf-Parkinson-White syndrome; ICD-9: 426.7; ICD-10: I45.6), cardiomegaly (which incorporates Athlete’s heart; ICD-9: 429.3; ICD-10: I51.7), congenital heart disease (ICD-9: 745-746; ICD-10: Q20-Q24), and coarctation of the aorta (ICD-9: 747.1; ICD-10: Q25.1); hypertension (ICD-10: I10-I16 and I1A) was added as a post-hoc exploratory analysis. The presence or absence of the following systemic conditions was recorded: sarcoidosis (ICD-9: 135; ICD-10: D86), glycogen storage disease (ICD-9: 271.0; ICD-10: E74.0), Fabry (-Anderson) disease (ICD-9: 272.7; ICD-10: E75.21), hemochromatosis (ICD-9: 275.01, 275.02, 275.03; ICD-10: E83.11), amyloidosis (ICD-9: 277.3; ICD-10: E85), and Friedreich ataxia (ICD-9: 334.0; ICD-10: G11.11). However, no patients with Friedreich ataxia were identified within the cohorts based on the ICD-9/ICD-10 codes.

### Model inference

The evaluated AI model was version 1.0 of the Viz HCM device, which was developed separately to this manuscript using proprietary methods. This device initially assessed the ECG data for the appropriate format to determine if the source file was supported and contained the essential components that are required to inspect the ECG metadata and signal quality. It then inspected the metadata of the ECG source file to assess eligibility of the patient ECG data for analysis. It also inspected the ECG signal for the presence of a pacemaker, in which case it would not provide an interpretation. It then performed a quality check of the ECG signal to identify ECGs with a sub-optimal signal (i.e., corrupt or missing leads). It then pre-processed and converted the ECG signal into different tensors, which were fed into an ensemble of neural network models for inference.

The device output consisted of an HCM classification score between 0 and 1 (with scores closer to 1 indicating a greater likelihood of HCM) and a binary output (HCM suspected or not suspected) above a sponsor-defined threshold. It also outputted the inference time. The model was installed at MGB for use in this study and received only the deidentified ECG waveform file.

### Statistical analysis

The statistical analysis was performed in R (version 4.0.2). The predefined primary endpoints were the sensitivity and specificity for suspected HCM detection. The predefined secondary endpoint was the positive predictive value (PPV). The predefined secondary assessments included the area under the receiving operating characteristic curve (AUC), negative predictive value (NPV) and descriptive summaries of the model inference time. They also included subgroup analyses of sensitivity of HCM characterization (i.e., non-obstructive versus obstructive), and sensitivity and specificity for patient demographics (age range, sex, race, ethnicity), technical parameters (ECG machine model), hospital site, presence of co-existing ECG findings or abnormalities, and presence of chart-based co-existing conditions. Post-hoc exploratory analyses included subgroup analyses of sensitivity based on patient sarcomere variant status, left ventricular wall thickness and left ventricular ejection fraction. The patient demographic and technical parameter data were derived from clinical databases and ECG file metadata respectively; any missing data were treated as “Unknown” and no data were imputed.

The confidence intervals (CIs) for sensitivity and specificity were calculated using the Agresti-Coull method which allows sensitivities and specificities to be less than 0% or greater than 100%. The CIs for PPV, NPV and AUC were calculated using bootstrapping with 2,000 intervals. Formal comparison testing was not performed between subgroups with the 95% CIs being the focus for evaluating any differences. The sample sizes were powered based on preliminary model results to achieve a sensitivity and specificity that would enable PPV ≥ 5.0% using prevalence of 0.002 (1 in 500).

## Results

### Cohort selection

The cohort selection process yielded 300 HCM-Positive and 3,000 HCM-Negative cases (Figure 1). The subsequent ECG annotations showed these cases inadvertently included 7 (2.3%) HCM-Positive and 88 (2.9%) HCM-Negative cases with paced rhythm and/or corrupt lead(s), and they were removed from the cohort. The model also identified 2 (0.7%) HCM-Positive and 7 (0.2%) HCM-Negative cases that were non-compliant based on its quality control checks. Model inference was performed on the remaining 291 HCM-Positive and 2905 HCM-Negative cases.

The HCM-Positive cases included 149 (51.2%) non-obstructive and 142 (48.8%) obstructive HCM cases (Table 1). These cases came from patients with mean age (± standard deviation; SD) of 61.3 ± 16.8 years and from 139 (47.8%) female patients; 15 (5.2%) patients had a sarcomere variant, 52 (17.9%) had no sarcomere variant and 224 (77.0%) had an unknown sarcomere variant status. The HCM-Negative cases came from patients with mean age (± SD) of 59.8 ± 18.0 years and from 1501 (51.7%) female patients. The race, ethnicity and ECG machine model baseline information for HCM-Positive and HCM-Negative cases is provided in Table 1.

**Table 1.** Baseline demographic and technical information for ECG cases.

|  | HCM-Positive |  |  | HCM-Negative |
| --- | --- | --- | --- | --- |
|  | Non-obstructive | Obstructive | Total |  |
| **Number of cases** | 149 | 142 | 291 | 2905 |
| **Sarcomere variant status** |  |  |  |  |
| Present | 10 (6.7%) | 5 (3.5%) | 15 (5.2%) | N/A |
| Absent | 24 (16.1%) | 28 (19.7%) | 52 (17.9%) | N/A |
| Unknown | 115 (77.2%) | 109 (76.8%) | 224 (77.0%) | N/A |
| **Gender** |  |  |  |  |
| Female | 61 (40.9%) | 78 (54.9%) | 139 (47.8%) | 1501 (51.7%) |
| Male | 88 (59.1%) | 64 (45.1%) | 152 (52.2%) | 1404 (48.3%) |
| **Age** |  |  |  |  |
| Mean ± SD (years) | 58.4 ± 18.0 | 64.2 ± 15.1 | 61.3 ± 16.8 | 59.8 ± 18.0 |
| <40 years | 27 (18.1%) | 11 (7.7%) | 38 (13.1%) | 486 (16.7%) |
| 40-65 years | 68 (45.6%) | 63 (44.4%) | 131 (45.0%) | 1178 (40.6%) |
| >65 years | 54 (36.2%) | 68 (47.9%) | 122 (41.9%) | 1241 (42.7%) |
| **Race** |  |  |  |  |
| American Indian or Alaska Native | 0 (0.0%) | 0 (0.0%) | 0 (0.0%) | 2 (0.1%) |
| Asian | 12 (8.1%) | 3 (2.1%) | 15 (5.2%) | 94 (3.2%) |
| Black or African American | 22 (14.8%) | 9 (6.3%) | 31 (10.7%) | 264 (9.1%) |
| Native Hawaiian or Other Pacific Islander | 0 (0.0%) | 0 (0.0%) | 0 (0.0%) | 1 (0.0%) |
| White | 109 (73.2%) | 114 (80.3%) | 223 (76.6%) | 2243 (77.2%) |
| Other | 3 (2.0%) | 6 (4.2%) | 9 (3.1%) | 193 (6.6%) |
| Two or more | 2 (1.3%) | 2 (1.4%) | 4 (1.4%) | 21 (0.7%) |
| Declined | 1 (0.7%) | 3 (2.1%) | 4 (1.4%) | 34 (1.2%) |
| Unknown | 0 (0.0%) | 5 (3.5%) | 5 (1.7%) | 53 (1.8%) |
| **Ethnicity** |  |  |  |  |
| Hispanic | 3 (2.0%) | 4 (2.8%) | 7 (2.4%) | 170 (5.9%) |
| Non-Hispanic | 126 (84.6%) | 120 (84.5%) | 246 (84.5%) | 2506 (86.3%) |
| Declined | 6 (4.0%) | 11 (7.7%) | 17 (5.8%) | 120 (4.1%) |
| Unknown | 14 (9.4%) | 7 (4.9%) | 21 (7.2%) | 109 (3.8%) |
| **ECG machine model** |  |  |  |  |
| CASE | 0 (0.0%) | 0 (0.0%) | 0 (0.0%) | 2 (0.1%) |
| D3K | 17 (11.4%) | 12 (8.5%) | 29 (10.0%) | 542 (18.7%) |
| MAC16 | 0 (0.0%) | 0 (0.0%) | 0 (0.0%) | 3 (0.1%) |
| MAC2K | 10 (6.7%) | 17 (12.0%) | 27 (9.3%) | 165 (5.7%) |
| MAC55 | 116 (77.9%) | 108 (76.1%) | 224 (77.0%) | 2028 (69.8%) |
| MAC5K | 0 (0.0%) | 0 (0.0%) | 0 (0.0%) | 1 (0.0%) |
| MV360 | 1 (0.7%) | 0 (0.0%) | 1 (0.3%) | 37 (1.3%) |
| S8500 | 5 (3.4%) | 5 (3.5%) | 10 (3.4%) | 127 (4.4%) |

### Model performance

The model identified HCM with sensitivity 68.4% (95% CI: 62.8 to 73.5%; 199 true positive cases and 92 false negative cases) and specificity 99.1% (95% CI: 98.7 to 99.4%; 2880 true negative cases and 25 false positive cases; Table 2). Assuming a prevalence of 1:500, these results corresponded to PPV 13.7% (95% CI: 10.1 to 19.9%) and NPV 99.9% (95% CI: 99.9 to 99.9%). The model had AUC 0.975 (95% CI: 0.965 to 0.982; Figure 2). There were no notable differences in sensitivity (a difference was considered notable when the 95% CIs did not overlap) for obstructive versus non-obstructive HCM, sarcomere variant status, left ventricular wall thickness and left ventricular ejection fraction. The model had mean processing time (± SD) of 1.8 ± 0.3 seconds; the range was 0.8 to 7.8 seconds. Example ECGs for different model accuracies are provided in Figure 3.

**Figure 2.**
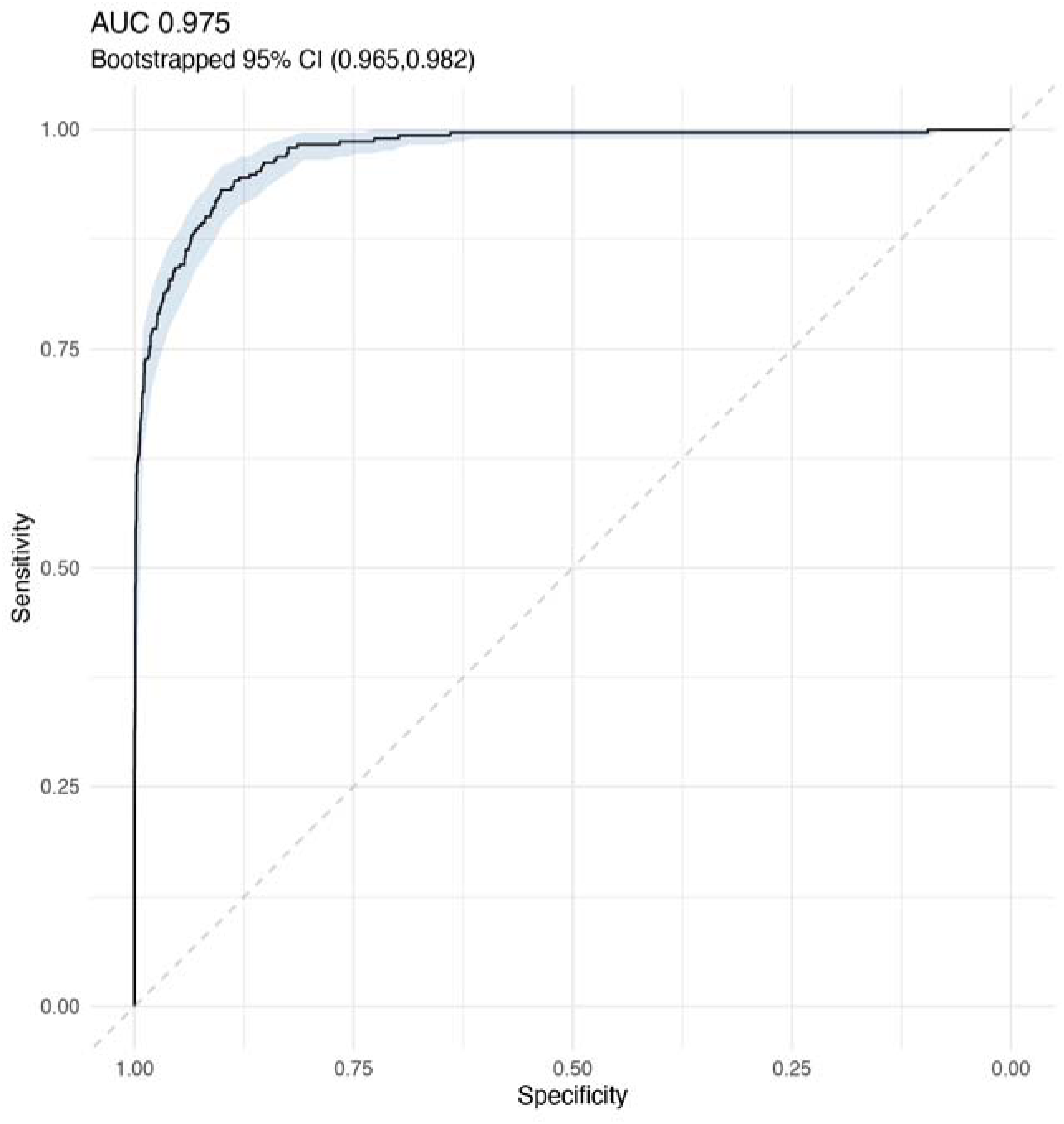
Model performance. This receiver operating characteristic curve includes the 95% CI in shading.

**Figure 3.**
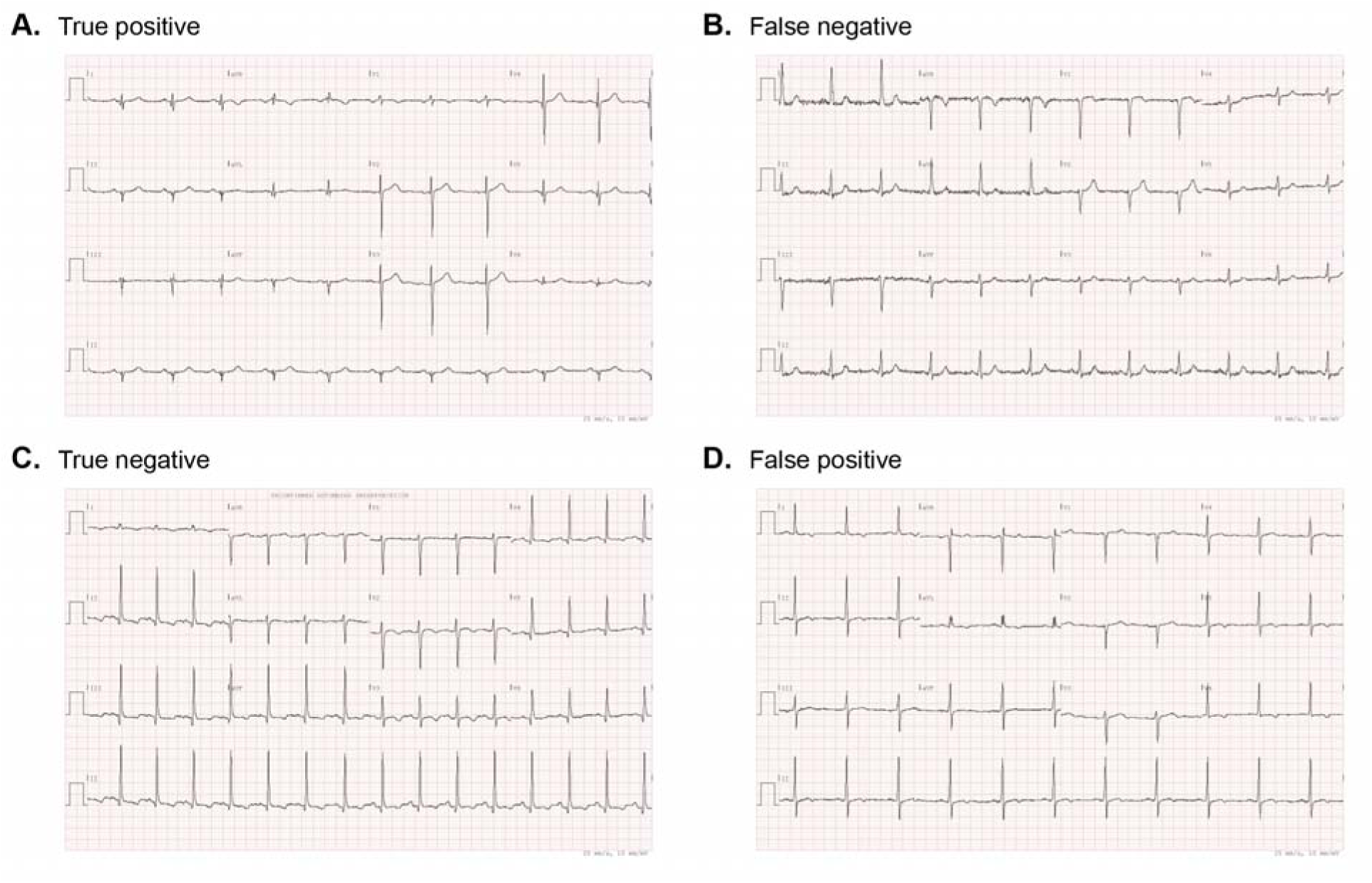
Example ECG cases. These cases demonstrate HCM-Positive cases that the model correctly identified as suspected HCM (A) or did not correctly identify (B), and HCM-Negative cases that the model correctly identified as not suspected (C) or did not correctly identify (D).

**Table 2.** Model performance summary. This summary includes the 95% CIs in parentheses. There were 11 cases without a reported left ventricular wall thickness and 3 cases without a reported left ventricular ejection fraction.

| **Overall** |  |
| --- | --- |
| AUC | 0.975 (0.965 to 0.982) |
| Sensitivity | 68.4% (62.8 to 73.5%) |
| Specificity | 99.1% (98.7 to 99.4%) |
| PPV | 13.7% (10.1 to 19.9%) |
| NPV | 99.9% (99.9 to 99.9%) |
| **Sensitivity for subgroups** |  |
| *HCM characterization* |  |
| Non-obstructive cases (N = 149) | 68.5% (60.6 to 75.4%) |
| Obstructive cases (N = 142) | 68.3% (60.2 to 75.4%) |
| *Sarcomere variant status* |  |
| Present (N = 15) | 73.3% (47.6 to 89.5%) |
| Absent (N = 52) | 75.0% (61.7 to 84.9%) |
| Unknown (N = 224) | 66.5% (60.1 to 72.4%) |
| *Left ventricular wall thickness* |  |
| <13mm (N = 47) | 74.5% (60.4 to 84.9%) |
| ≥13mm (N = 233) | 67.4% (61.1 to 73.1%) |
| *Left ventricular ejection fraction* |  |
| <50% (N = 10) | 60.0% (31.2 to 83.3%) |
| ≥50% (N = 278) | 68.7% (63.0 to 73.9%) |

### Demographic and technical subgroup performance

The model identified HCM with AUC greater than 0.970 for all gender, age, race, ethnicity, ECG machine model and hospital site subgroups (Supplementary Table 1) except for the unknown ethnicity subgroup (AUC 0.964) and hospital site 3 (AUC 0.948). There were no demographic or technical subgroups where there was a notable difference in sensitivity or specificity (when the 95% CIs from subgroups did not overlap). However, the model did show a trend for lower sensitivity in cases from female patients (61.2% [95% CI: 52.8 to 68.9%]) compared with male patients (75.0% [95% CI: 67.5 to 81.2%]), yet higher specificity (99.5% [95% CI: 99.0 to 99.8%] in cases from female patients versus 98.7% [95% CI: 98.0 to 99.2%] from male patients). The model also showed a trend for higher sensitivity for cases from patients aged <40 years (84.2% [95% CI: 69.2 to 92.9%]) compared with patients aged 40 to 65 years (64.1% [95% CI: 55.6 to 71.8%]) or >65 years (68.0% [95% CI: 59.3 to 75.7%]). Among the three most common racial groups in the cohort, the model had a trend for higher sensitivity for cases from Asian patients (73.3% [95% CI: 47.6 to 89.5%]) compared with Black or African American patients (64.5% [95% CI: 46.9 to 79.0%]) and White patients (66.8% [95% CI: 60.4 to 72.7%]); the specificities were similar across these three racial groups (98.9% [95% CI: 93.6 to 100.4%] for cases from Asian patients, 99.2% [95% CI: 97.1 to 100.0%] from Black or African American patients, and 99.1% [95% CI: 98.6 to 99.4%] from White patients).

### Other ECG findings subgroup performance

The model was also assessed for its ability to identify HCM in the presence or absence of other ECG findings (Supplementary Table 2). For many ECG findings, the performance was similar in the presence or absence of each finding. The findings where there was a notable difference in sensitivity (when the 95% CIs did not overlap) included atrial fibrillation (46.2% [95% CI: 28.7 to 64.5%] when present versus 70.6% [95% CI: 64.8% to 75.7%] when absent), left ventricular hypertrophy (74.7% [95% CI: 68.7 to 79.9%] when present versus 45.2% [95% CI: 33.4 to 57.5%] when absent), non-specific ST segment change(s) (76.0% [95% CI: 68.5 to 82.2%] when present versus 60.3% [95% CI: 52.0 to 68.0%] when absent), no pathologic finding (25.0% [95% CI: 6.3 to 59.9%] when present versus 69.6% [95% CI: 64.0 to 74.7%] when absent). The findings where there was a notable difference in specificity including left ventricular hypertrophy (95.8% [95% CI: 92.6 to 97.7%] when present versus 99.5% [95% CI: 99.1 to 99.7%] when absent) and non-specific ST segment change(s) (97.7% [95% CI: 95.5 to 98.8%] when present versus 99.4% [95% CI: 99.0 to 99.6%] when absent).

### Presence of comorbid conditions subgroup performance

The model was assessed for its ability to identify HCM in the presence or absence of comorbid conditions including cardiopulmonary and systemic conditions (Supplementary Table 3). For many comorbid conditions, the performance was similar in the presence or absence of each finding. There were no comorbid conditions with a notable difference in sensitivity (when the 95% CIs did not overlap). The comorbid conditions where there was a notable difference in specificity included nonrheumatic aortic stenosis (96.2% [95% CI: 93.1 to 98.0%] when present versus 99.4% [95% CI: 99.0 to 99.7%] when absent) and congenital heart disease (97.1% [95% CI: 93.1 to 98.9%] when present versus 99.3% [95% CI: 98.9 to 99.5%] when absent).

## Discussion

This study assessed the ability of an AI model to identify people with suspected HCM based on their ECG. This model overall performed well, achieving a sensitivity of 68.4% and specificity of 99.1%. These metrics corresponded to a PPV of 13.7%, NPV of 99.9% and AUC of 0.975. Based on these results, this model was granted a de novo request by the FDA as the first device under regulation number 21 CFR 870.2380 for cardiovascular machine learning-based notification software (10).

A key consideration for the current algorithm was ensuring an adequate balance between identifying true positive and false positive cases as reflected by the PPV. The operating point for this device was therefore chosen to have a higher specificity compared with sensitivity. Two algorithms in the research setting have demonstrated a similar AUC; they have utilized operating points that balance sensitivity and specificity differently, which we note would correspond to a lower PPV. An algorithm developed at the Mayo Clinic performed with AUC 0.96, sensitivity 87% and specificity 90% (15). A separate algorithm developed at Yale New Haven Hospital had AUC 0.96, sensitivity 91% and specificity 91% (16).

The clinical utility of an ECG-based diagnostic device in initial screening for HCM is that it can be implemented at lower cost and complexity than echocardiography-based assessment. The specific application may vary from one setting to another. An example is the large-scale screening of young, mostly healthy individuals prior to participation in physical activities such as athletes and military recruits; the sensitivity of 84.2% amongst the <40 years subgroup is particularly appealing in this setting. Given the ability to function automatically, the device could separately analyze all ECGs obtained in defined clinical settings to identify patients who could benefit from cardiology evaluation.

The device demonstrated broadly consistent performance across different demographic groups. A notable difference was the trend for higher sensitivity amongst ECGs obtained from male compared with female patients, which may relate to the larger voltages typically seen on ECGs from male patients. This study tested the current FDA-approved version of this device, which utilizes an “operating point” that is akin to a normal threshold value for a laboratory test. Future device iterations may consider a lower operating point for women.

The race of patients is known to impact their ECG (17). This study had included over 100 ECG cases (∼10% of which were HCM-Positive) from patients from each of Asian, Black or African American, and White racial backgrounds. The sensitivity was similar between patients from the Black or African American and White subgroups, while patients from the Asian subgroup had a trend for improved sensitivity. The cause for this increased sensitivity is not apparent, although echocardiographic differences including more frequent apical hypertrophy have been described in Asian compared to European healthcare centers (18). The consistency across these racial groups provides reassurance about device generalizability. Its ongoing ability to perform for people from different racial subgroups, especially those with smaller representation in this cohort, should continue to be assessed.

A limitation of this study included that it was performed in a retrospective manner on cases where a diagnosis of HCM had already been made, while its potential use is in a prospective manner on cases where a diagnosis of HCM has not been made. This restriction was in large part because this assessment of the AI model aimed to show that it has the accuracy and therefore potential to be beneficial. The key clinical effectiveness question moving forward will be how many cases of undiagnosed HCM it helps identify. Further assessments of its clinical effectiveness, as well as its performance in settings beyond those tested as part of this assessment, will be required.

The study design required HCM-Positive cases to demonstrate ECG criteria for left ventricular hypertrophy during initial selection. ECGs were assessed again for left ventricular hypertrophy at the time of ECG annotations, which was one of 16 different ECG findings. During these ECG annotations, a proportion of HCM-Positive cases were annotated as not containing left ventricular hypertrophy (i.e., were HCM-Positive left ventricular hypertrophy-absent). In a review of such cases, many appeared to have borderline left ventricular hypertrophy. We were reassured that the model still performed with sensitivity of 45.2% in cases with this discrepancy. Similarly, we were reassured that the model maintained specificity of 95.8% on the HCM-Negative left ventricular hypertrophy-present cases, showing that it has the ability to discriminate between the presence or absence of HCM in spite of the presence of left ventricular hypertrophy.

## Conclusion

The ability to detect HCM on 12-lead ECG has the potential to assist with screening for HCM. We demonstrated the accuracy of an AI model in identifying suspected HCM on ECG including its performance across a range of demographic and technical subgroups, and in the presence of other ECG findings or comorbid conditions.

## Supporting information

STARD Checklist

## Data Availability

All data produced in the present study are available upon reasonable request to the authors.

## Acknowledgements

The authors thank the broader Mass General Brigham AI and Viz.ai teams for their assistance with this project.

## Tables

**Supplementary Table 1.** Model performance across demographic and technical subgroups. This table includes the 95% confidence intervals in parentheses.

|  | HCM-Positive<br>N | HCM-Negative<br>N | AUC | Sensitivity | Specificity |
| --- | --- | --- | --- | --- | --- |
| **Gender** |  |  |  |  |  |
| Female | 139 | 1501 | 0.979<br>(0.971 to 0.985) | 61.2%<br>(52.8 to 68.9%) | 99.5%<br>(99.0 to 99.8%) |
| Male | 152 | 1404 | 0.971<br>(0.954 to 0.984) | 75.0%<br>(67.5 to 81.2%) | 98.7%<br>(98.0 to 99.2%) |
| **Age** |  |  |  |  |  |
| <40 years | 38 | 486 | 0.977<br>(0.932 to 1.000) | 84.2%<br>(69.2 to 92.9%) | 99.8%<br>(98.7 to 100.1%) |
| 40 to 65 years | 131 | 1178 | 0.973<br>(0.963 to 0.982) | 64.1%<br>(55.6 to 71.8%) | 99.0%<br>(98.2 to 99.4%) |
| >65 years | 122 | 1241 | 0.973<br>(0.959 to 0.983) | 68.0%<br>(59.3 to 75.7%) | 99.0%<br>(98.3 to 99.5%) |
| **Race** |  |  |  |  |  |
| American Indian<br>or Alaska Native | 0 | 2 | - | - | 100.0%<br>(29.0 to 105.2%) |
| Asian | 15 | 94 | 0.987<br>(0.962 to 0.999) | 73.3%<br>(47.6 to 89.5%) | 98.9%<br>(93.6 to 100.4%) |
| Black or African<br>American | 31 | 264 | 0.981<br>(0.961 to 0.994) | 64.5%<br>(46.9 to 79.0%) | 99.2%<br>(97.1 to 100.0%) |
| Native Hawaiian<br>or Other Pacific<br>Islander | 0 | 1 | - | - | 100.0%<br>(16.7 to 103.9%) |
| White | 223 | 2243 | 0.972<br>(0.959 to 0.981) | 66.8%<br>(60.4 to 72.7%) | 99.1%<br>(98.6 to 99.4%) |
| Other | 9 | 193 | 1.000<br>(0.997 to 1.000) | 100.0%<br>(65.5 to 104.5%) | 99.5%<br>(96.8 to 100.2%) |
| Two or more | 4 | 21 | 1.000<br>(1.000 to 1.000) | 75.0%<br>(28.9 to 96.6%) | 100.0%<br>(81.8 to 102.8%) |
| Declined | 4 | 34 | 1.000<br>(0.956 to 1.000) | 75.0%<br>(28.9 to 96.6%) | 100.0%<br>(87.9 to 101.9%) |
| Unknown | 5 | 53 | 0.996<br>(0.966 to 1.000) | 80.0%<br>(36.0 to 98.0%) | 100.0%<br>(91.9 to 101.3%) |
| **Ethnicity** |  |  |  |  |  |
| Hispanic | 7 | 170 | 1.000<br>(0.995 to 1.000) | 100.0%<br>(59.6 to 105.0%) | 98.2%<br>(94.7 to 99.6%) |
| Non-Hispanic | 246 | 2506 | 0.973<br>(0.962 to 0.982) | 67.5%<br>(61.4 to 73.0%) | 99.2%<br>(98.7 to 99.5%) |
| Declined | 17 | 120 | 0.989 | 64.7% | 100.0% |
|  |  |  | (0.964 to 1.000) | (41.2 to 82.8%) | (96.3 to 100.6%) |
| Unknown | 21 | 109 | 0.964<br>(0.923 to 0.990) | 71.4%<br>(49.8 to 86.4%) | 99.1%<br>(94.5 to 100.3%) |
| **ECG machine model** |  |  |  |  |  |
| CASE | 0 | 2 | - | - | 100.0%<br>(29.0 to 105.2%) |
| D3K | 29 | 542 | 0.990<br>(0.981 to 0.996) | 62.1%<br>(43.9 to 77.4%) | 99.4%<br>(98.3 to 99.9%) |
| MAC16 | 0 | 3 | - | - | 100.0%<br>(38.3 to 105.6%) |
| MAC2K | 27 | 165 | 0.974<br>(0.940 to 0.994) | 81.5%<br>(62.8 to 92.3%) | 98.8%<br>(95.4 to 99.9%) |
| MAC55 | 224 | 2028 | 0.972<br>(0.960 to 0.981) | 67.9%<br>(61.5 to 73.6%) | 99.0%<br>(98.5 to 99.4%) |
| MAC5K | 0 | 1 | - | - | 100.0%<br>(16.7 to 103.9%) |
| MV360 | 1 | 37 | 1.000<br>(1.000 to 1.000) | 100.0%<br>(16.7 to 103.9%) | 100.0%<br>(88.8 to 101.8%) |
| S8500 | 10 | 127 | 0.980<br>(0.949 to 0.997) | 60.0%<br>(31.2 to 83.3%) | 100.0%<br>(96.5 to 100.6%) |
| **Hospital Sites** |  |  |  |  |  |
| Site 1 | 111 | 968 | 0.982<br>(0.971 to 0.990) | 73.9%<br>(65.0 to 81.2%) | 98.9%<br>(98.0 to 99.4%) |
| Site 2 | 107 | 978 | 0.986<br>(0.980 to 0.992) | 70.1%<br>(60.8 to 78.0%) | 99.6%<br>(98.9 to 99.9%) |
| Site 3 | 73 | 959 | 0.948<br>(0.915 to 0.970) | 57.5%<br>(46.1 to 68.2%) | 99.0%<br>(98.1 to 99.5%) |

**Supplementary Table 2.**
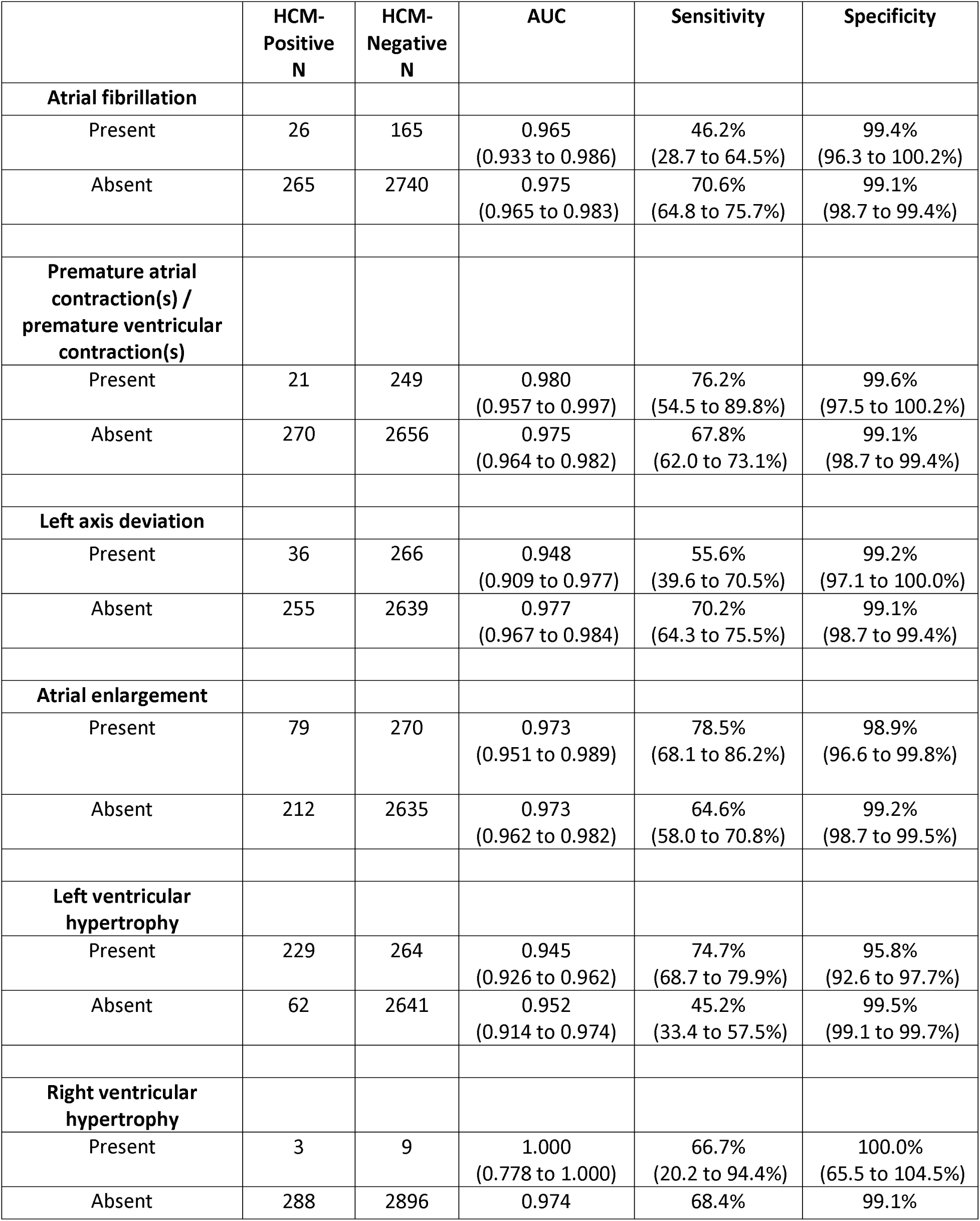
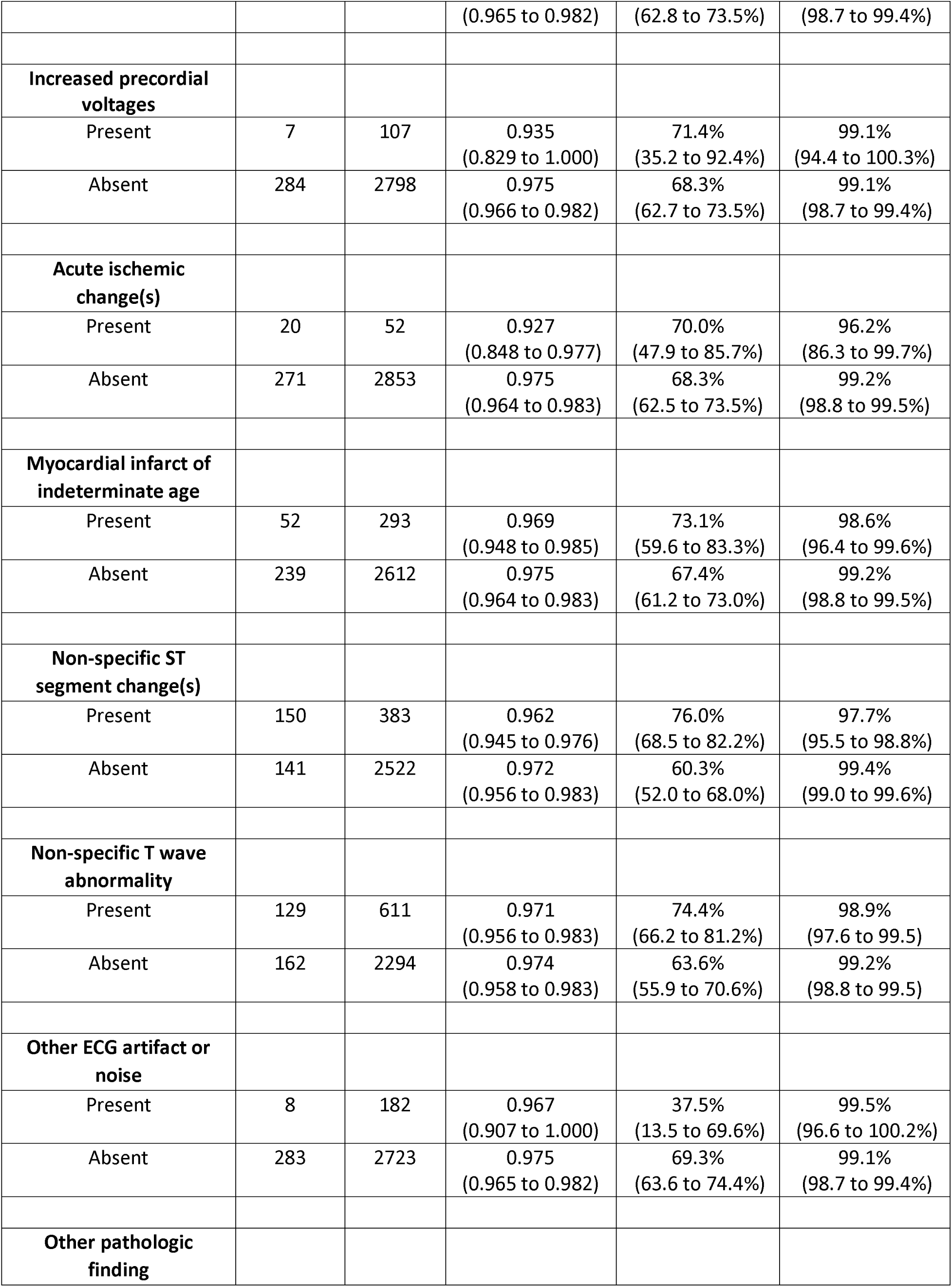
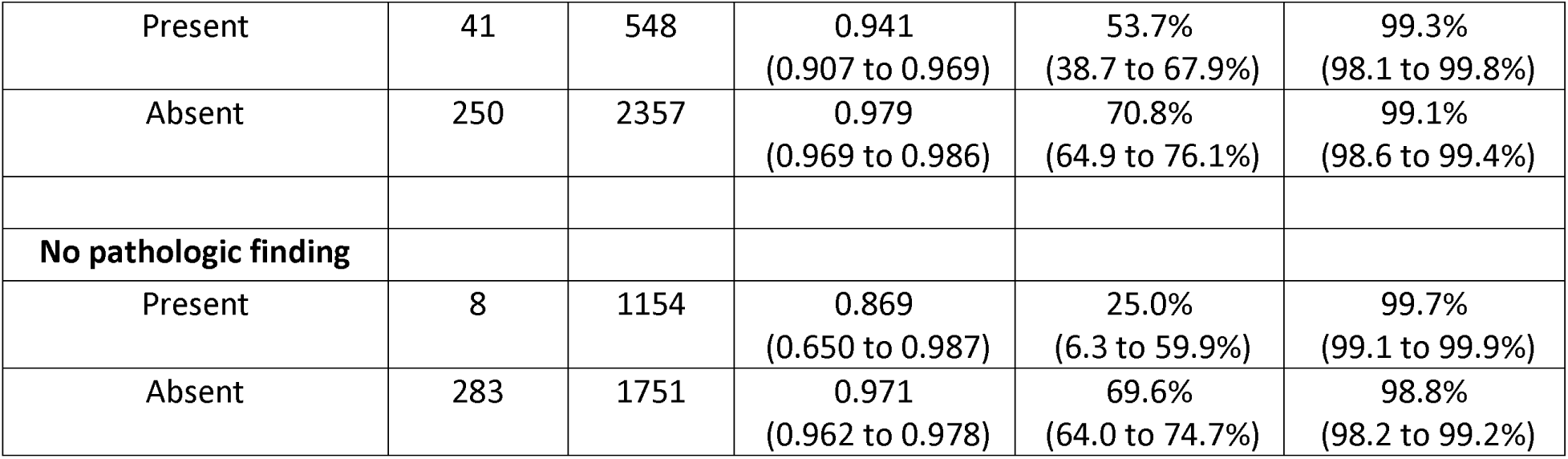
Model performance in the presence of other ECG findings. The presence of other ECG findings was determined from ECG review by cardiologists. This table includes the 95% confidence intervals in parentheses.

**Supplementary Table 3.** Model performance in the presence of comorbid conditions. The presence of comorbid conditions was determined based on the presence of ICD-9 or ICD-10 codes in a patient’s clinical record. This table includes the 95% confidence intervals in parentheses. There were 28 HCM-Negative cases for which the patients did not have ICD-9 or ICD-10 codes available.

|  | HCM-Positive<br>N | HCM-Negative<br>N | AUC | Sensitivity | Specificity |
| --- | --- | --- | --- | --- | --- |
| **Hypertension** |  |  |  |  |  |
| Present | 193 | 1881 | 0.967<br>(0.957 to 0.976) | 64.8%<br>(57.8 to 71.2%) | 98.9%<br>(98.4 to 99.3%) |
| Absent | 98 | 996 | 0.984<br>(0.963 to 0.996) | 75.5%<br>(66.1 to 83.0%) | 99.5%<br>(98.8 to 99.8%) |
| **Ischemic heart disease** |  |  |  |  |  |
| Present | 125 | 1340 | 0.974<br>(0.964 to 0.983) | 70.4%<br>(61.9 to 77.7%) | 98.7%<br>(97.9 to 99.2%) |
| Absent | 166 | 1537 | 0.977<br>(0.963 to 0.987) | 66.9%<br>(59.4 to 73.6%) | 99.5%<br>(99.0 to 99.8%) |
| **Pulmonary hypertension** |  |  |  |  |  |
| Present | 22 | 158 | 0.964<br>(0.926 to 0.989) | 68.2%<br>(47.1 to 83.8%) | 98.7%<br>(95.2 to 99.9%) |
| Absent | 269 | 2719 | 0.975<br>(0.965 to 0.983) | 68.4%<br>(62.6 to 73.7%) | 99.2%<br>(98.7 to 99.4%) |
| **Pulmonary heart disease** |  |  |  |  |  |
| Present | 7 | 87 | 1.000<br>(1.000 to 1.000) | 100.0%<br>(59.6 to 105.0%) | 100.0%<br>(94.9 to 100.8%) |
| Absent | 284 | 2790 | 0.974<br>(0.964 to 0.981) | 67.6%<br>(62.0 to 72.8%) | 99.1%<br>(98.7 to 99.4%) |
| **Nonrheumatic aortic stenosis** |  |  |  |  |  |
| Present | 42 | 265 | 0.955<br>(0.927 to 0.975) | 69.0%<br>(53.9 to 81.0%) | 96.2%<br>(93.1 to 98.0%) |
| Absent | 249 | 2612 | 0.976<br>(0.966 to 0.984) | 68.3%<br>(62.2 to 73.7%) | 99.4%<br>(99.0 to 99.7%) |
| **Left bundle branch block** |  |  |  |  |  |
| Present | 26 | 113 | 0.896<br>(0.829 to 0.950) | 69.2%<br>(49.9 to 83.7%) | 96.5%<br>(91.0 to 98.9%) |
| Absent | 265 | 2764 | 0.977 | 68.3% | 99.2% |
|  |  |  | (0.967 to 0.984) | (62.5 to 73.6%) | (98.8 to 99.5%) |
| **Right bundle branch block** |  |  |  |  |  |
| Present | 16 | 233 | 0.945<br>(0.882 to 0.989) | 62.5%<br>(38.5 to 81.6%) | 98.3%<br>(95.5 to 99.5%) |
| Absent | 275 | 2644 | 0.976<br>(0.967 to 0.983) | 68.7%<br>(63.0 to 73.9%) | 99.2%<br>(98.8 to 99.5%) |
| **Pre-excitation syndrome** |  |  |  |  |  |
| Present | 4 | 14 | 1.000<br>(1.000 to 1.000) | 75.0%<br>(28.9 to 96.6%) | 100.0%<br>(74.9 to 103.6%) |
| Absent | 287 | 2863 | 0.974<br>(0.965 to 0.981) | 68.3%<br>(62.7 to 73.4%) | 99.1%<br>(98.7 to 99.4%) |
| **Cardiomegaly** |  |  |  |  |  |
| Present | 131 | 312 | 0.961<br>(0.944 to 0.976) | 66.4%<br>(57.9 to 73.9%) | 98.1%<br>(95.8 to 99.2%) |
| Absent | 160 | 2565 | 0.976<br>(0.960 to 0.986) | 70.0%<br>(62.5 to 76.6%) | 99.3%<br>(98.8 to 99.5%) |
| **Congenital heart disease** |  |  |  |  |  |
| Present | 23 | 170 | 0.984<br>(0.964 to 0.996) | 69.6%<br>(48.9 to 84.6%) | 97.1%<br>(93.1 to 98.9%) |
| Absent | 268 | 2707 | 0.974<br>(0.963 to 0.982) | 68.3%<br>(62.5 to 73.6%) | 99.3%<br>(98.9 to 99.5%) |
| **Coarctation of the aorta** |  |  |  |  |  |
| Present | 0 | 7 | - | - | 85.7%<br>(46.7 to 99.5%) |
| Absent | 291 | 2870 | 0.975<br>(0.965 to 0.982) | 68.4%<br>(62.8 to 73.5%) | 99.2%<br>(98.8 to 99.4%) |
| **Sarcoidosis** |  |  |  |  |  |
| Present | 1 | 29 | 1.000<br>(1.000 to 1.000) | 100.0%<br>(16.7 to 103.9%) | 96.6%<br>(81.4 to 100.8%) |
| Absent | 290 | 2848 | 0.975<br>(0.965 to 0.982) | 68.3%<br>(62.7 to 73.4%) | 99.2%<br>(98.7 to 99.4%) |
| **Glycogen storage disease** |  |  |  |  |  |
| Present | 0 | 1 | - | - | 100.0%<br>(16.7 to 103.9%) |
| Absent | 291 | 2876 | 0.974 | 68.4% | 99.1% |
|  |  |  | (0.964 to 0.982) | (62.8 to 73.5%) | (98.7 to 99.4%) |
| **Fabry (-Anderson) disease** |  |  |  |  |  |
| Present | 0 | 9 | - | - | 88.9%<br>(54.3 to 100.2%) |
| Absent | 291 | 2868 | 0.974<br>(0.965 to 0.982) | 68.4%<br>(62.8 to 73.5%) | 99.2%<br>(98.8 to 99.4%) |
| **Hemochromatosis** |  |  |  |  |  |
| Present | 0 | 18 | - | - | 100.0%<br>(79.3 to 103.1%) |
| Absent | 291 | 2859 | 0.974<br>(0.964 to 0.982) | 68.4%<br>(62.8 to 73.5%) | 99.1%<br>(98.7 to 99.4%) |
| **Amyloidosis** |  |  |  |  |  |
| Present | 1 | 26 | 1.000<br>(1.000 to 1.000) | 100.0%<br>(16.7 to 103.9%) | 100.0%<br>(84.8 to 102.4%) |
| Absent | 290 | 2851 | 0.974<br>(0.965 to 0.982) | 68.3%<br>(62.7 to 73.4%) | 99.1%<br>(98.7 to 99.4%) |

